# Non-prescription Plant-Based Aphrodisiacs Inducing Nephritis and Nephrosis in sub-Saharan African Men: A Systematic Review Protocol

**DOI:** 10.1101/2025.09.16.25335902

**Authors:** Senzelokuhle M. Nkabini, Lindiwe Nkabini-Anderson, Fezile Zuma

## Abstract

**Background:** Sub-Saharan African (SSA) men’s erotic desires for sexual prowess, stamina, and sturdy penile strength have created a ‘multi-million-dollar industry’ for businesses retailing non-prescription plant-based aphrodisiacs (NPBA’s). Globally, the registered and approved NPBA mass-producing sexual enhancement supplements (SES’s) retailing industry, has reached almost $300 million in 2025, and is projected to increase by 6.9% yearly up until 2035. The gross increase will be approximately $600 million. Unlike the global market that heavily relies on manufactured SES’s, a vast majority of male SSA citizens still prefer natural NPBA’s. These NPBA’s are natural plant species (herbs, flowering plants & roots) that are either consumed in their natural form or mixed with warm water to extract the plants or roots properties. Currently in SSA, almost 600 plant species have been discovered in less than 10 countries, and these plants are currently being used as NPBA’s by men. However, due to the SSA region comprising of 49 countries and approximately 1.6 billion citizens, a large majority of untested, and unapproved plant species are currently being used as NPBA’s. This is dangerous because the global, continental (Africa), and regional (SSA) health & medical science communities have not tested the toxicity levels of these NPBA’s. This lack of knowledge has led to health risks such as nephritis (inflammation of the kidneys), and nephrosis (excessive protein loss due to damaged kidney filters) in SSA men. Moreover, the easy access in obtaining NPBA’s in SSA and the excessive usage of them by men, has led to a spike in chronic kidney diseases (CKD) such as nephritis and nephrosis. CKD is currently a growing public health concern in SSA that has received minimal attention due to it being a non-communicable illness. Approximately 174 million SSA citizens are affected by CKD which has caused 77,000 deaths in the region, and the severity of this issue is proven by CKD increasing by 0,85% annually. Currently, the excessive use of unapproved NPBA’s in SSA has been included as one of the leading causes of CKD in men.

**Methods:** The primary aim of this systematic review is to map out and synthesise evidence of, NPBA’s inducing nephritis and nephrosis in SSA men from existing literature. The following databases will be utilized to search for studies: PubMed, PsycINFO, ProQuest, ERIC (Education Resources Information Center), Cochrane Reviews, WHO, and Scopus. The Preferred Reporting Items for Systematic and Meta Analyses (PRISMA) ScR flow chart/diagram presented in figure 1 will be utilized to summarize the study selection process.

**Conclusion:** Despite CKD/Kidney disease/nephritis & nephrosis not being included as: (i) a part of the NCD’s that cause a large number of deaths globally, (ii) the 4 major NCD’s, that are reported on within several global WHO NCD publications, within Africa and the SSA region this chronic disease is a serious public health crisis. Unfortunately, untested, unapproved, and unregulated NPBA’s that currently exist in SSA, are also suspected of predominantly contributing to this crisis in men. Moreover, throughout the African continent and SSA region, only three countries (Ethiopia, South Africa & Zambia) offer both renal replacement by transplantation, and renal replacement therapy by dialysis. Hence, the escalating death rates caused by CKD/Kidney disease/nephritis & nephrosis. The proposed systematic review will generate findings pertaining to NPBA’s inducing nephritis and nephrosis in SSA men. These findings will/can reveal the current existing literature gaps regarding NPBA’s, CKD, nephritis and nephrosis.

**Systematic reviews registration:** PROSPERO (CRD420251146160)

## Background

Sub-Saharan African (SSA) men’s erotic desires for sexual prowess, stamina, and sturdy penile strength have created a ‘multi-million-dollar industry’ for businesses retailing non-prescription plant-based aphrodisiacs (NPBA’s). Globally, the registered and approved NPBA mass-producing sexual enhancement supplements (SES’s) retailing industry, has reached almost $300 million in 2025, and is projected to increase by 6.9% yearly up until 2035 (1). The gross increase will be approximately $600 million (1). Unlike the global market that heavily relies on manufactured SES’s, a vast majority of male SSA citizens still prefer natural NPBA’s (2). These NPBA’s are natural plant species (herbs, flowering plants & roots) that are either consumed in their natural form or mixed with warm water to extract the plants or roots properties (2,3). Currently in SSA, almost 600 plant species have been discovered in less than 10 countries, and these plants are currently being used as NPBA’s by men (4,5). However, due to the SSA region comprising of 49 countries and approximately 1.6 billion citizens, a large majority of untested, and unapproved plant species are currently being used as NPBA’s. This is dangerous because the global, continental (Africa), and regional (SSA) health & medical science communities have not tested the toxicity levels of these NPBA’s. This lack of knowledge has led to health risks such as nephritis (inflammation of the kidneys), and nephrosis (excessive protein loss due to damaged kidney filters) in SSA men.

Within the SSA region, both past and present, NPBA’s have occasionally been used for creating interest in sexual intercourse, and maintaining sexual enjoyment throughout the human ageing process (6,7,8,9). Due to these aphrodisiacs being plant-based, and a part of traditional medicines that have been used to heal various ailments they are either orally consumed, swallowed or administered through the rectum with an enema in their natural state by men (6,7,8,9). NPBA’s have the natural advantage of being assumed to be safe, healthy and a ‘much-needed’ eliminator of most chronic diseases. This is because of their vegetative form that has not been distorted, and transformed into a tablet/syrup that has a high volume of chemical preservatives to prevent it from decomposing. However, despite the infamousness of NPBA’s being a natural resource that assists with sexual dysfunctions in SSA, they have not been tested for any other side-effects that they might cause to the human body (6,7,8,9). Moreover, the easy access in obtaining NPBA’s in SSA and the excessive usage of them by men, has led to a spike in chronic kidney diseases (CKD) such as nephritis and nephrosis (6,7,8,9). CKD is currently a growing public health concern in SSA that has received minimal attention due to it being a non-communicable illness (10). Approximately 174 million SSA citizens are affected by CKD which has caused 77,000 deaths in the region, and the severity of this issue is proven by CKD increasing by 0,85% annually (10). Currently, the excessive use of unapproved NPBA’s in SSA has been included as one of the leading causes of CKD in men.

The World Health Organisation (WHO) initially created, *The Global Action Plan for The Prevention and Control of Non-Communicable Diseases (NCD’s) 2013-2020* in order to decrease the 36 million global deaths (63%) caused by NCD’s (11). However, this 2013-2020 action plan was only intended for addressing NCD’s such as cardiovascular diseases, all forms of cancers, chronic respiratory illnesses, and diabetes (11). These aforementioned NCD’s were responsible for 86% of deaths in lower-middle-income countries (LMIC’s), and SSA has approximately 41 countries out of 49 that are defined as LMIC’s (11). Towards the end of 2013-2020 the *Draft Implementation Road Map 2023-2030 for The Global Action Plan for The Prevention and Control of Non-Communicable Diseases 2013-2030* was drafted (12). The 2023-2030 action plan focuses on: (i) accelerating national response to NCD’s, (ii) prioritising the implementation of interventions to NCD’s, and (iii) ensuring reliable national data on NCD’s risk factors (12). In spite of this, within the 2023-2030 action plan, the section focusing on its *scope, purpose, and modalities* does not feature CKD/Kidney disease/nephritis & nephrosis as part of the ‘4 by 4 NCD agenda’ or the ‘5 by 5 NCD agenda’ (12). Moreover, the WHO NCD data portal does not feature CKD/Kidney disease/nephritis & nephrosis as part of the NCD’s that cause a large number of deaths globally (10,13). This is in spite of the WHO NCD portal featuring diseases and risk factors such as harmful alcohol use, smoking, cancer, diabetes, and diabetes (13). These diseases and risk factors are one of the many lead causes of CKD/Kidney disease/nephritis & nephrosis. Furthermore, CKD/Kidney disease/nephritis & nephrosis has not been included in the 4 major NCD’s, that are reported on within several global WHO NCD publications (11,14,15,16,17). Nonetheless, this chronic disease does feature in *WHO-African Region: Communicable and Non-Communicable Diseases in Africa in 2021/2022*, but this subtlety denotes CKD/Kidney disease/nephritis & nephrosis as an ‘African disease’ instead of being designated as a global public health crisis (37). Hence, the relevance and necessity of studies such as this systematic review.

NPBA’s inducing nephritis and nephrosis in SSA men is part of the global challenges, that the United Nations (UN) wants to address through the 17 sustainable development goals (SDGs) (18). NPBA’s inducing nephritis and nephrosis in SSA men is included as part of the challenges that should be addressed by SDG 3: *Good Health and Well-being*. Due to the severity of preventing and controlling NCD’s, WHO has created a sub-section for this SDG titled *SDG Target 3.4 Non-communicable Diseases and Mental Health*. This sub-section focuses on practical interventions that work towards providing treatment, in order to prevent premature mortality and reduce death rates that are linked to NCD’s (19). 18 million people under the age of 70 have died due to untreated NCD’s, despite 62% of these premature deaths being preventable (19). *SDG 3*, *SDG Target 3.4*, and the *Draft Implementation Road Map 2023-2030 for The Global Action Plan for The Prevention and Control of Non-Communicable Diseases 2013-2030* are notable programmes for dealing with NCD’s. However, the lack in recognition of CKD/Kidney disease/nephritis & nephrosis, has created a serious gap in research and knowledge regarding this NCD.

Evidence of NPBA’s inducing nephritis and nephrosis in SSA men will be systematically mapped, synthesised, and literature gaps will be exposed through the guidance of primary and secondary research questions. The primary research question that will guide this review is: What factors contribute to SSA men utilizing non-prescribed plant-based aphrodisiacs? The secondary research questions are: What are the experiences of SSA men that use non-prescription plant-based aphrodisiacs? What evidence exists regarding non-prescription plant-based aphrodisiacs inducing nephritis and nephrosis in SSA men? Databases such as PubMed, Cochrane Reviews, and EBSCOhost were utilized to conduct a preliminary database search in order to prevent the duplication of studies. Based on the research teams (SMN, LN-A & FZ) knowledge there has been no other systematic review conducted on NPBA’s inducing nephritis, and nephrosis in SSA men. However, there is a study titled *Natural aphrodisiacs consumption by male workers in the former Katanga province, DR Congo* (8). The results from this study found no evidence of nephrotoxicity or hepatotoxicity caused by the consumption of natural aphrodisiacs. The results from this study can be attributed to the investigation only obtaining data from a small sample (research participants), that is located in one province, and within one SSA country. Thus, further research is required hence the results from this systematic review will assist organisations such as WHO, CKD-Africa, International Society of Nephrology, Kidney Disease Improving Global Outcome (KIDIGO), World Kidney Day, National Kidney Foundation, Kidney Research United Kingdom, National Kidney Federation, National Kidney Foundation of South Africa (NKFSA), and the International Society of Nephrology (ISN).

## Methods

The primary aim of this systematic review is to map out and synthesise evidence of, NPBA’s inducing nephritis and nephrosis in SSA men from existing literature. All forms of studies, grey literature and peer-reviewed journal articles focusing on non-prescription plant-based aphrodisiacs, and nephritis & nephrosis in SSA men will be sourced. The primary research question that will guide this review is: What factors contribute to SSA men utilizing non-prescribed plant-based aphrodisiacs? The secondary research questions are: What are the experiences of SSA men that use non-prescription plant-based aphrodisiacs? What evidence exists regarding non-prescription plant-based aphrodisiacs inducing nephritis and nephrosis in SSA men? The P.I.Co (population, interest, context) framework was utilized to address the eligibility, and adequateness of the primary and secondary research questions. This framework (P.I.Co) has been adapted from the P.I.C.O framework, which is used for quantitative systematic reviews (20). However, due to some systematic reviews focusing on qualitative components, the P.I.Co framework was created (20).

### P.I.Co (Population, Interest, Context) framework

#### Population

Non-prescription plant-based aphrodisiacs are herbs, flowering plants, and roots utilised to stimulate sexual desire, improve sexual function, as well as enhancing sexual performance (21,22).

Age: 19+ years old is the average age range that is allowed to use commercially produced SES’s. However, due to NPBA’s being untested, unregulated, and retailed with no prescription requirement, the age range might be lower. Furthermore, the age range is also dependant on the practiced socio-cultural rituals of each SSA country, and when communities in these SSA countries choose to usher boys into manhood, and teach them about the man’s role in: society, in his family, and in the marital bedroom (23,24,25).

#### Interest

Nephritis and nephrosis in men, is the inflammation, and overall damage of the kidneys blood vessels which later leads to: blood being present in the urine, high blood pressure, foamy urine, weight gain due to fluid retention, and swelling (around the eyes, feet & ankles) (26,27).

#### Context

SSA is a region within the African continent, that has 49 countries (28).

#### Sources of evidence

All studies, grey and empirical literature containing evidence on NPBA’s that induce nephritis and nephrosis in SSA men.

#### Publication Year Range

01/01/2016-30/09/2025 Language: No language restriction

### Information sources and search strategy

All searches will be conducted by SMN, LN-A and FZ for studies and study designs published in peer-reviewed journals, grey literature, published and unpublished dissertations, case studies, reviews, essays, theses and symposium abstracts. The following databases will be utilized to search for studies: PubMed, PsycINFO, ProQuest, ERIC, Cochrane Reviews, WHO, and Scopus. The search strategy will use the Boolean term ‘OR’ to separate words or search terms. The following search terms will be utilised: non-prescription OR plant-based OR aphrodisiac OR inducing OR nephritis OR nephrosis OR sub-Saharan African/ SSA OR men. A preliminary data base search was conducted by the research team using the search terms, and the results are presented in Table 1. The preliminary search produced all studies, qualitative and quantitative full-text grey literature and peer-reviewed literature with no language restrictions, and published within the search time-line from 01January 2016 till 30 September 2025.

**Table 1:**
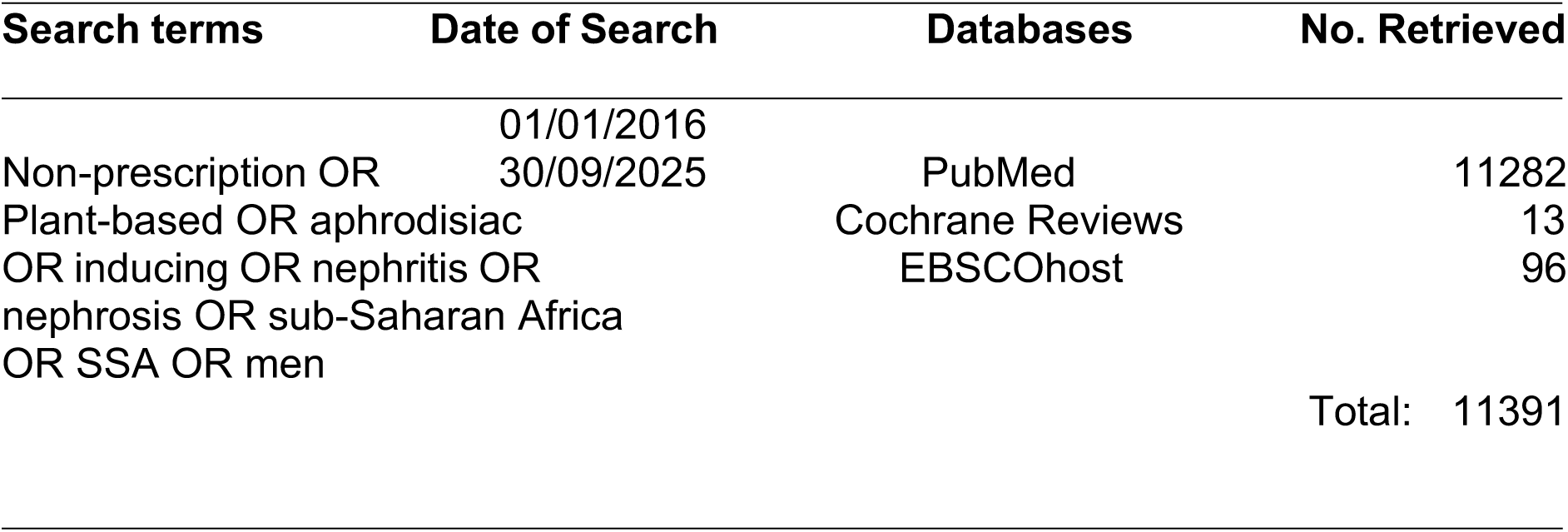
Preliminary database search results.

**Table 2:** Data charting table.

|  |
| --- |
| Author and year of publication |
| Article title |
| Study aim |
| Country |
| Study design/methodology |
| Study setting |
| Study population |
| Duration of the study |
| Factors contributing to NPBA's inducing nephritis & nephrosis in SSA men |
| The experiences of SSA men that use NPBA's |
| NPBA's inducing nephritis and nephrosis in SSA men |
| Population age |
| Relevant/significant findings |
| Key conclusions of article |
| Notes |

### Inclusion and exclusion criteria

The selection of eligible studies will be based on the title (NPBA’s inducing nephritis and nephrosis), abstract, setting (SSA), study population (SSA men) and the findings in full-text studies, books, grey literature and journal articles relating to NPBA’s, and nephritis & nephrosis in SSA men. All database searches will be conducted on a weekly basis by SMN, LN-A and FZ to ensure new literature is included. Only studies that adhere to the following criteria will be included: (i) NPBA’s, and nephritis & nephrosis in SSA men, (ii) qualitative and quantitative study designs iii) studies and articles published from 2016 to 2025 and, (iv) only studies, books, grey literature or journal articles that are restricted to human ages 19+ years old, full-text, have references/citations and have been peer-reviewed. Studies that are: (i) published before 2016, will be excluded.

### Data management and study selection

The framework, enhancements to the framework and guidelines provided by Arksey and O’Malley’s (29), Levac *et al* (30), Daudt *et al* (31), and Johanna Briggs (32) will guide this review. The Preferred Reporting Items for Systematic and Meta Analyses (PRISMA) ScR flow chart/diagram presented in figure 1 will be utilized to summarize the study selection process (33,34). The authors and the two research assistants will ensure that all retrieved literature will be exported and saved to an Endnote 21.3 library folder, in order to enable the study to be reproduced for a second time at a later stage. Moreover, this process will allow SMN, LN-A and FZ to create separate libraries for each database, in order to import references, remove duplicates, and organize the findings.

**Figure 1:**
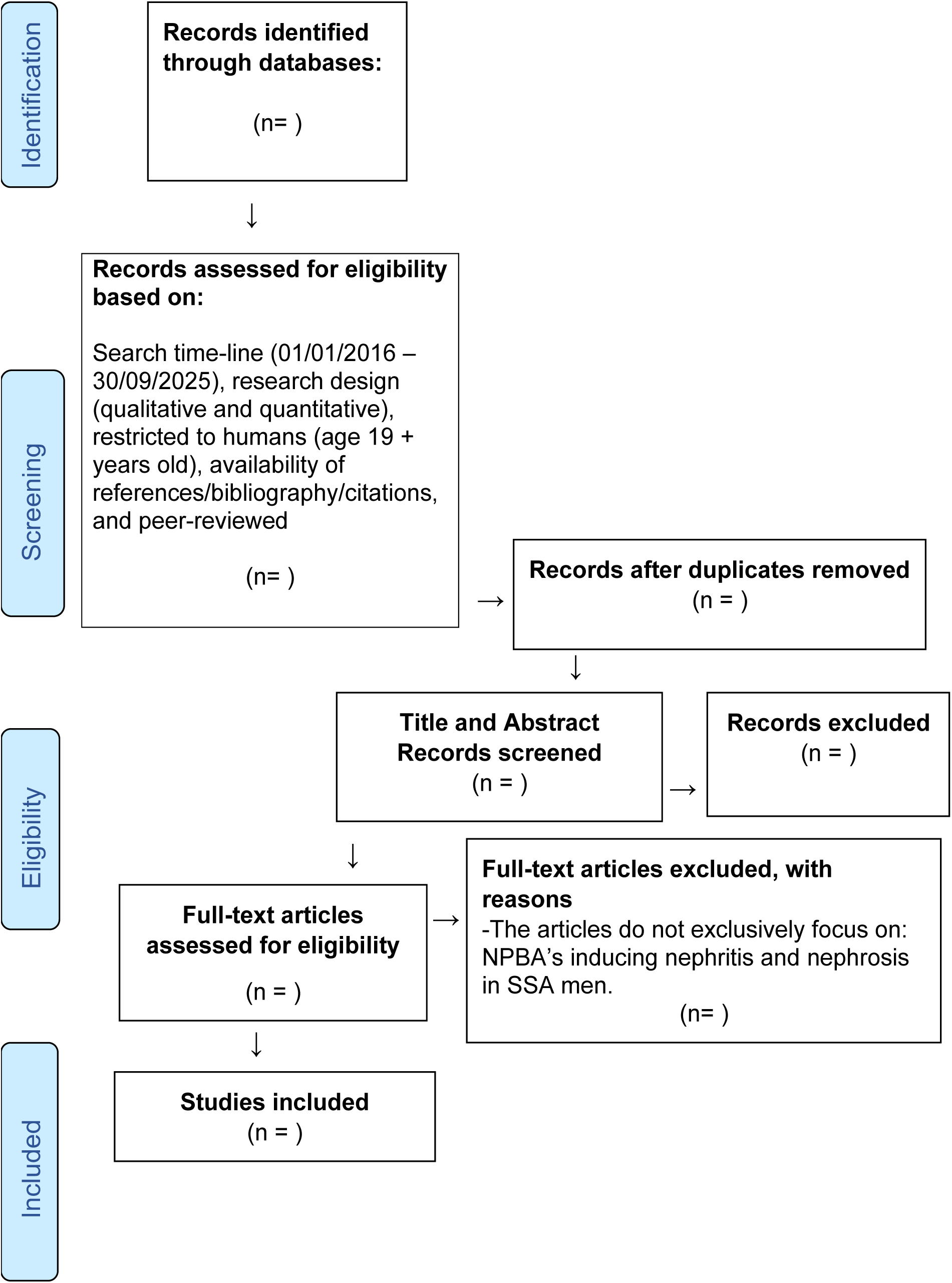
The Preferred Reporting Items for Systematic and Meta Analyses (PRISMA) ScR flow chart/diagram

### Data extraction

A data charting table will be used in order to document the extracted data. Studies that meet the inclusion criteria will be included on the charting form. The use of the NVivo data analysis software and Braun and Clarke’s (35) thematic framework will guide the qualitative analysis. The data extraction form will be updated by SMN, LN-A and FZ on a weekly basis to guarantee accurateness.

### Risk of bias assessment

Studies or articles that only feature few countries that are located in the SSA region will not be included, because the research results from these studies do not represent SSA as a whole. In order to avoid biasness the review will utilise the mixed-method appraisal tool (MMAT) version 2018 to appraise the quality of all included evidence (36). SMN, LN-A & FZ will be responsible for assigning ratings of 100% for high average sources, 50% average and 25% for low-quality articles.

### Discrepancies between the protocol and the systematic review

Discrepancies between the protocol, the actual review and the reasons and consequences thereof will be reported in the final report.

### Data synthesis

A narrative synthesis will be conducted, which will provide texts and tables in order to synthesize and discuss the data of the studies and the methods, as previously described in the data extraction section.

## Results

The results of this systematic review will be disseminated through publication in peer-reviewed scientific journals. The data will also be made available to humanitarian organisations such as WHO, CKD-Africa, International Society of Nephrology, Kidney Disease Improving Global Outcome (KIDIGO), World Kidney Day, National Kidney Foundation, Kidney Research United Kingdom, National Kidney Federation, National Kidney Foundation of South Africa (NKFSA), and the International Society of Nephrology (ISN). The findings from this study will inform the future research projects that these organisations embark on.

## Discussion

Despite CKD/Kidney disease/nephritis & nephrosis not being included as: (i) a part of the NCD’s that cause a large number of deaths globally, (ii) the 4 major NCD’s, that are reported on within several global WHO NCD publications, within Africa and the SSA region this chronic disease is a serious public health crisis (10,13, 11,14,15,16,17). Unfortunately, untested, unapproved, and unregulated NPBA’s that currently exist in SSA, are also suspected of predominantly contributing to this crisis in men. Moreover, throughout the African continent and SSA region, only three countries (Ethiopia, South Africa & Zambia) offer both renal replacement by transplantation, and renal replacement therapy by dialysis (37). Hence, the escalating death rates caused by CKD/Kidney disease/nephritis & nephrosis. The proposed systematic review will generate findings pertaining to NPBA’s inducing nephritis and nephrosis in SSA men. These findings will/can reveal the current existing literature gaps regarding NPBA’s, CKD, nephritis and nephrosis. Humanitarian organisations such as WHO, CKD-Africa, International Society of Nephrology, Kidney Disease Improving Global Outcome (KIDIGO), World Kidney Day, National Kidney Foundation, Kidney Research United Kingdom, National Kidney Federation, National Kidney Foundation of South Africa (NKFSA), and the International Society of Nephrology (ISN) can conduct further research on this topic.

## Data Availability

All data produced in the present work are contained in the manuscript

## Acknowledgments

The authors express their gratitude to: the Faculty of Humanities at the University of Pretoria.

## Authors contributions

SMN, LN-A and FZ conducted the preliminary database search for the protocol, conceptualised, drafted, edited, reviewed, sent out the draft protocol manuscript for constructive feedback, and approved the final manuscript.

## Ethics approval and consent to participate

Not applicable.

## Consent for publication

Not applicable.

## Availability of data material

The required data is included in the manuscript.

## Abbreviations

CKD: Chronic Kidney Disease
ISN: International Society of Nephrology
KIDIGO: Kidney Disease Improving Global Outcome
LMIC: Lower-Middle-Income-Country
MMAT: Mixed-Method Appraisal Tool
NCD: Non-Communicable Disease
NKFSA: National Kidney Foundation of South Africa
NPBA: Non-prescription Plant-Based Aphrodisiacs
P.I.Co: Population. Interest. Context
PRISMA: Preferred Reporting Items for Systematic and Meta Analyses
SES: Sexual Enhancement Supplements
SSA: sub-Saharan Africa
SDG: Sustainable Development Goals
UN: United Nations
WHO: World Health Organisation

---

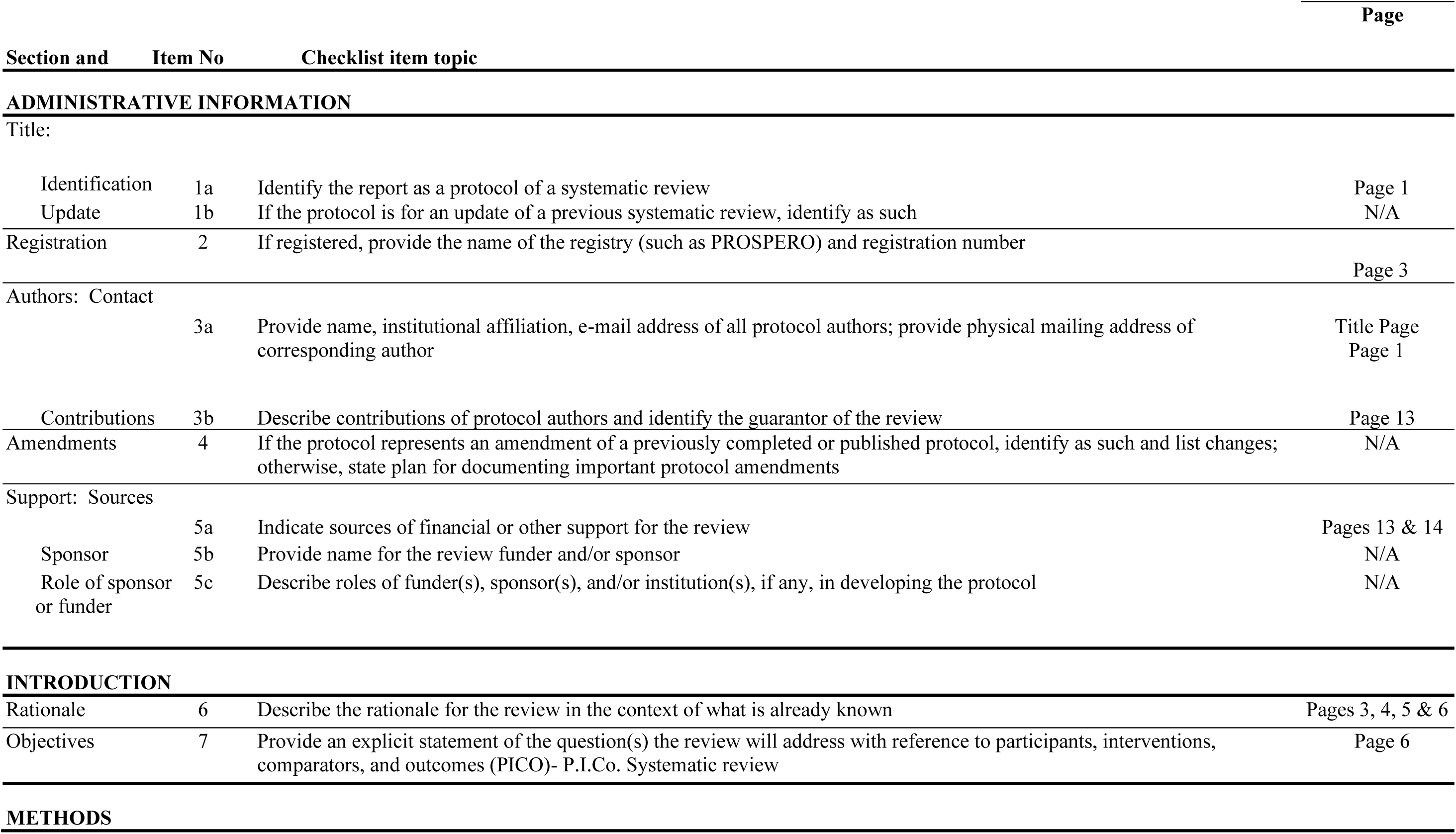
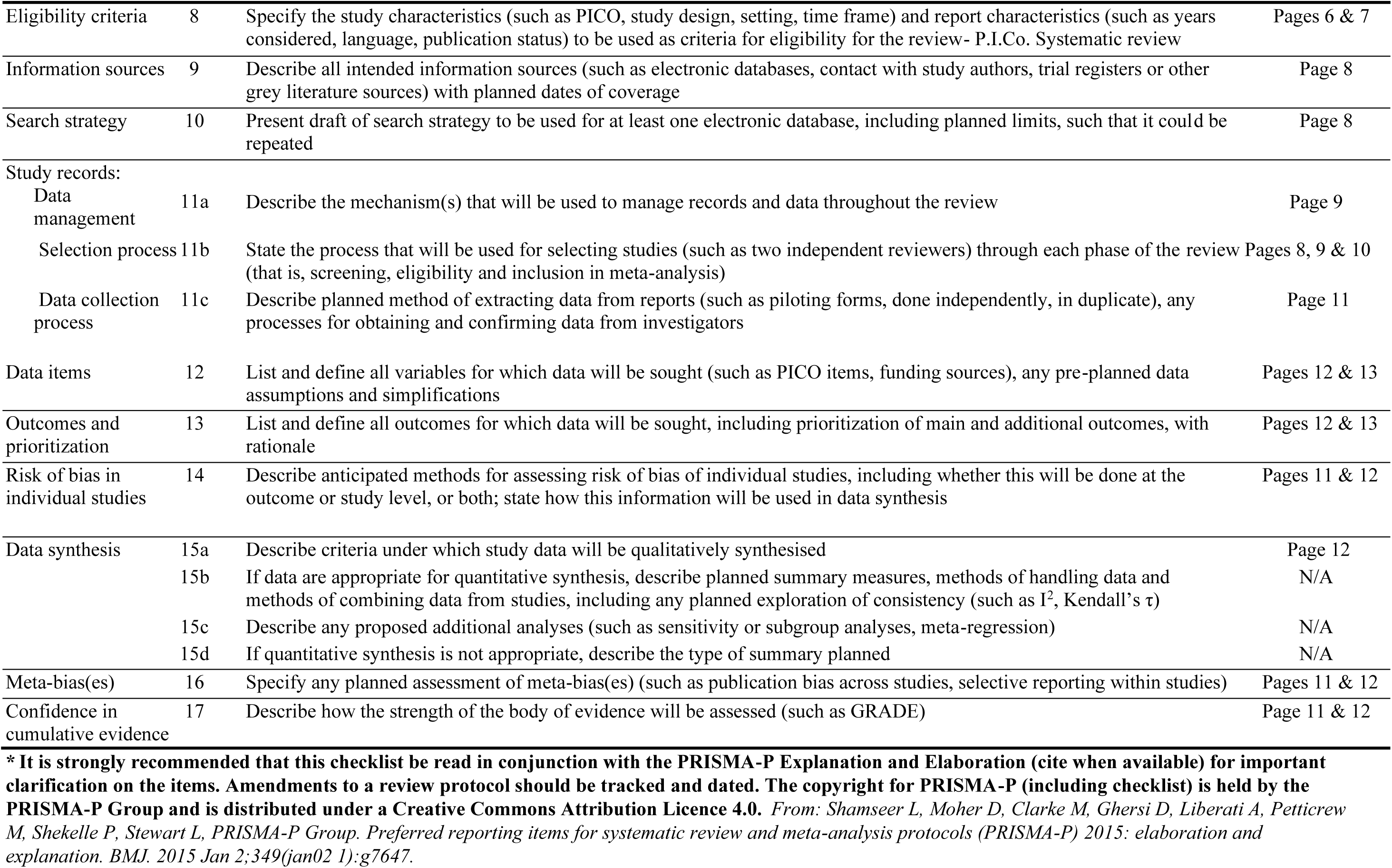
PRISMA-P (Preferred Reporting Items for Systematic review and Meta-Analysis Protocols) 2015 checklist: recommended items to address in a systematic review protocol*.

